# SARS-CoV-2 in wastewater from Mexico City used for irrigation in the Mezquital Valley: quantification and modelling of geographic dispersion

**DOI:** 10.1101/2021.06.07.21258522

**Authors:** Yaxk’in Coronado, Roberto Navarro, Carlos Mosqueda, Valeria Valenzuela, Juan Pablo Perez, Víctor González-Mendoza, Mayra de la Torre, Jorge Rocha

## Abstract

Quantification of SARS-CoV-2 in urban wastewaters has emerged as a cheap, efficient strategy to follow trends of active COVID-19 cases in populations. Moreover, mathematical models have been developed that allow prediction of active cases following the temporal patterns of viral loads in wastewaters. In Mexico, no systematic efforts have been reported in the use of this strategies. In this work, we quantified SARS-CoV-2 in rivers and irrigation canals in the Mezquital Valley, Hidalgo, an agricultural region where wastewater from Mexico City is distributed and used for irrigation. Using quantitative RT-PCR, we detected the virus in 6 out of 8 water samples from rivers, and 5 out of 8 water samples from irrigation canals. Notably, samples showed a general consistent trend of having the highest viral loads in the sites closer to Mexico City, indicating that this is the main source that contributes to detection. Using the data for SARS-CoV-2 concentration in the river samples, we generated a simplified transport model that describes the spatial patterns of dispersion of virus in the river. We suggest that this model can be extrapolated to other wastewater systems that require knowledge of spatial patterns of viral dispersion at a geographic scale. Our work highlights the need for improved practices and policies related to the use of wastewater for irrigation in Mexico and other countries.

## Introduction

The ongoing global pandemic of COVID-19 disease, caused by severe acute respiratory syndrome coronavirus 2 (SARS-CoV-2), is a public health emergency of international concern (Organization and Fund (UNICEF), 2020a, 2020b). SARS-CoV-2 ribonucleic acid (RNA) has been detected in feces from both symptomatic and asymptomatic patients (Chen et al., 2020; Holshue et al., 2020; Jiehao et al., 2020; Tang et al., 2020; W. Wang et al., 2020; Zhang et al., 2020) and in wastewater (Ahmed et al., 2020; Lodder and Husman, 2020; Medema et al., 2020). For this reason, quantification of SARS-CoV-2 ARN in wastewater has emerged as a cheap, efficient method for monitoring active cases in large populations (Ahmed *et al*., 2020; S. Wang *et al*., 2020), small towns (Kitajima *et al*., 2020; Randazzo *et al*., 2020), or campuses (Harris-Lovett *et al*., 2021). Notably, this strategy allows for a one-week anticipation in the active cases, compared to health systems registries, since asymptomatic individuals contribute to the viral load in wastewaters (Vallejo *et al*., 2020; Wu *et al*., 2020).

Mexico City has more than 661,446 confirmed cases of COVID-19 (Mexico City Government, 2020; consulted on June 1^st^, 2021); this is the city with the highest number of cases in the country. In Hidalgo, a state north of Mexican Valley Metropolitan Area (MVMA, Figure 1a), more than 39,012 positive cases have accumulated (State of Hidalgo Government, 2021), of which more than 11,000 cases (28%) correspond to the Mezquital Valley, a highly productive agricultural region.

**Figure 1.**
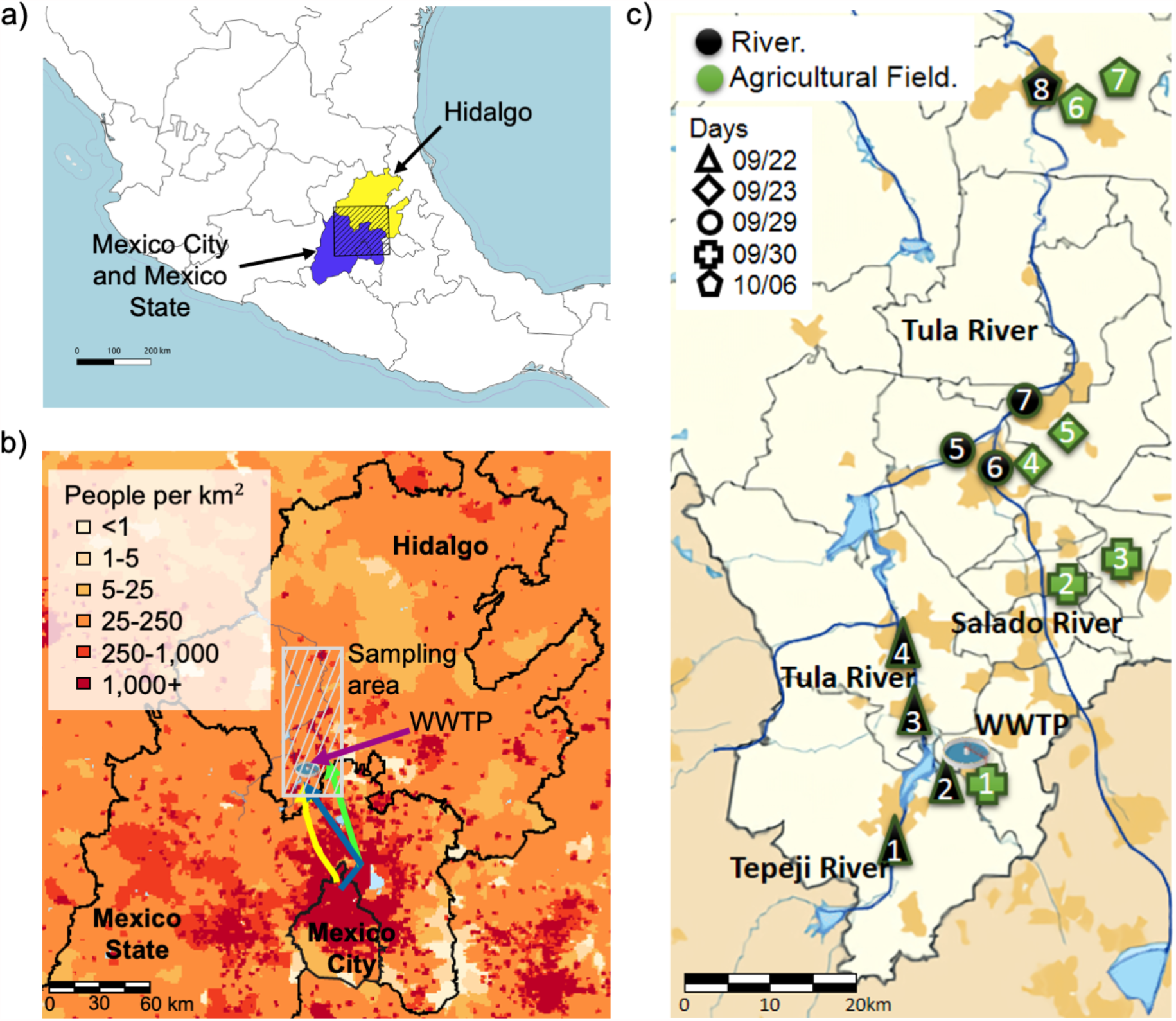
Study area. a) Location of the State of Hidalgo, in Mexico. Dashed area corresponds to Figure 1B. b) Mexico City is the main source of wastewater used for irrigation in the Mezquital Valley. HeatMap showing the population density in the study area. Yellow, blue and green arrows show the location and discharge of the main sewage systems. WWTP, Wastewater treatment plant. Dashed area corresponds to Figure 1c. c) Sampling locations and dates for samples in the Mezquital Valley. For agricultural field locations, samples of both water (irrigation canal) and soil (adjacent agricultural field) were collected.

To date, few systematic efforts have been made in Mexico to detect SARS-CoV-2 in wastewater. However, the Mezquital Valley has several relevant characteristics for the study of SARS-CoV-2 in wastewater: 1) agriculture production is maintained by using exclusively wastewater for irrigation (Contreras *et al*., 2017); 2) the wastewater source is the MVMA, the most populated metropolitan area worldwide, and the region that concentrates most active cases in Mexico (Figure 1b) (*Información referente a casos COVID-19 en México - datos*.*gob*.*mx/busca*); 3) the system includes one of the largest wastewater treatment facilities in Latin America, which feeds a river and a complex system of irrigation canals; 4) farmers, inhabitants and consumers in the Mezquital Valley are in contact with water, soil or agricultural products and 5) the use of wastewater for irrigation is one of the main causes that allowed that this Valley is no longer in extreme poverty (García-Salazar and García-Salazar, 2019).

Using data for SARS-CoV-2 concentration in wastewater, several models have been proposed for to find a correlation between the temporal patterns of viral concentration and the number of active COVID-19 cases (Hart and Halden, 2020b). Likewise, a previous work presented the first model of spatial and temporal patterns of viral loads in seaway systems, showing the dispersion of the virus in an urban region (Hart and Halden, 2020a). However, models for spatial patterns of viral dispersion are needed in open waterbodies that contain wastewater, in order to understand the virus transport and to identify the zones with higher risk of infection.

In this study we sampled water from the Tula River, Salado River and irrigation canals in the Mezquital Valley, which receive wastewater from Mexico City, to assess the presence of SARS-CoV-2 and generate a mathematical model that describes its spatial dispersion. We propose that our results highly relevant not only to follow the epidemic in Mexico City and municipalities in Hidalgo, but also to evaluate a possible risk of transmission through environmental matrices and measure the stability of SARS-CoV-2 with geographic resolution.

## Materials and Methods

### Sampling

Water samples were taken along the Tula River between the mouth of the Central Interceptor Tunnel and Ixmiquilpan; samples were also taken in the Salado River, which receives wastewater from the Grand Drainage Canal, and Tepeji river, which contains wastewaters from local municipalities (Figure 1). We also collected water samples from irrigation canals in locations representative of the Mezquital Valley. Appropriate safety equipment was always used, consisting of a cotton lab coat, latex gloves, KN95 respirator-mask, rubber boots, disposable cap, and safety glasses. A simple sampling technique was used (NOM, 1980; EPA, 2017), locating sites where the wastewater is well mixed near to the center of the flow channel, approximately between 40 and 60 percent of water depth, where turbulence is maximum, and there is minimum sedimentation of solids. For each location, three water samples were collected as follows: two samples of 400 ml in glass bottles, for SARS-CoV-2 detection and for microbiological analyses; and one 4 L sample in a plastic bottle, for physicochemical analysis. For each water sample collected at an irrigation canal, we sampled 500 g of soil from an adjacent agricultural field. All samples were immediately placed in ice until arrival at the laboratory. Water samples for SARS-CoV-2 detection were inactivated by incubation at 60°C for 1 h upon arrival at the laboratory.

### Physicochemical and Microbiological Analyses

Samples for physicochemical and microbiological analyses were stored in ice from sampling until delivery to external laboratories where determinations were carried out. Due to the remoteness of sampling sites, analyses were performed between 18 to 40 h after sampling. Water samples were analyzed for physicochemical parameters at *Laboratorio de Análisis Químicos Analíticos* (LAQA), at *Facultad de Química*, Universidad Autónoma de Querétaro (Querétaro, México). Physicochemical parameters measured were: Biochemical oxygen demand (BOD), chemical oxygen demand (COD); pH, total dissolved solids (TDS), suspended solids (SS), total suspended solids (TSS) and total solids (TS). Microbiological analysis of total coliforms (TC), fecal coliforms (FC), *Escherichia coli* and *Salmonella* (PCR detection) were analyzed (according to norm NOM-210-SSA-2014) at *Laboratorio para la Evaluación y Control de Riesgos Microbianos en Alimentos* (LECRIMA) at UAQ (Querétaro, México). The ‘most probable number’ method was used for quantification of TC, FC and *E. coli*.

### Viral RNA Extraction

For soil samples, viral particles were first eluted from 15 g of sample in 150 ml of 10% beef extract buffer (pH 7) by stirring for 30 min (Horm *et al*., 2012). Next, soil suspension and inactivated wastewater samples (150 ml) were filtered through a No. 5 Whatman filter (Millipore), and then through 0.22 µm polyethersulfone membrane (Corning). Next, viral particles were precipitated by adding NaCl (100 g/L) and Polyethilenglicol (22 g/L) and stirring for 20 min. Then, 105 ml samples were centrifuged at 12,000 x g for 2 hours, and the resulting pellet was suspended in 400 µl RNAse free water. Viral nucleic acids were extracted from these samples using the Purelink Viral DNA/RNA kit (Invitrogen). Purified nucleic acid samples were quantified by spectrophotometry in a Nanodrop One (Thermo Scientific).

### Quantification of SARS-CoV-2 RNA

For detection of SARS-CoV-2, the kit Decov2 Triplex (Genes2Life, Irapuato, Mexico) was used, which includes reverse transcription and PCR detection of three targets of N ORF (N1-FAM, N2-HEX and N3-TexasRed) in a single multiplex reaction. For each reaction, 5 µl of viral RNA were added as template in a total volume of 25 µl. The RT-qPCR reaction was performed in a QuantStudio 5 (Thermo Scientific) instrument, and settings for passive reference was changed to ‘none’. Duplicate SARS-CoV-2 detection reactions were performed for each RNA. For absolute quantification of SARS-CoV-2 in environmental samples, a standard curve was obtained using synthetic RNA targets provided with the detection kit. For all RNA samples, we included an internal control reaction for the detection of Pepper Mild Mottle Virus (PMMV), using the primers Fwd 5’-GAG TGG TTT GAC CTT AAC GTT TGA-3’, Rev 5’-TTG TCG GTT GCA ATG CAA GT-3’ and probe 5’-FAM-CCT ACC GAA GCA AAT G-BHQ1-3 (Zhang *et al*., 2005; Haramoto *et al*., 2020).

### Modelling of water dispersion of SARS-CoV-2 in the Tula River

The transport model for dispersion of virus in unsaturated media (Torkzaban *et al*., 2006) was adapted. The original model describes the viral concentration with respect to the distance from the virus input site and the decrease over time, and considers terms of adsorption and detachment of virus with respect to the soil-water interface and air-water interface. We considered only dispersion in the water bulk; we also considered that dispersion of virus is independent on time, and it depends on the distance. Hence, the attachment and detachment rate coefficients and the inactivation rate coefficient in soil and air media were equal to zero (κ^a^_att_ = 0, κ^a^_det_ = 0, κ^s^_att_= 0, κ^s^_det_ = 0, μ_s_ = 0 and μ_a_ = 0). We also assume that Darcy flux (q) and inactivation rate coefficient (μ_w_) are constant along the river. The variables amount of water (θ) and coefficient of dispersion (D) were fitted to the experimental data. Finally, we excluded the contributions of the adsorption from the general model and considered two entrances that affect the viral concentration. Detailed derivation of the equations is shown in Supplementary File 1. The resulting equation (1) is shown below:

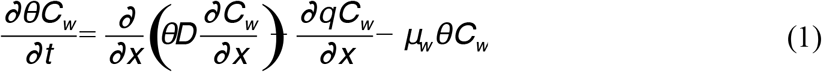

Where Cw, is the viral concentration and the other variables are previously decribed.

The equation was solved numerically with initial and boundary conditions using the software package Scipy Odeint in Python. The concentration values for the virus in the Tula River were obtained experimentally as described above.

## Results

### Social, agronomic and economic considerations of the study system

The Mezquital Valley is located 80 km north of Mexico City (Fig. 1). It is inhabited mainly by the indigenous *Otomí* group that speaks a native language called *hñähñu*. The average annual precipitation is 409 mm (Moreno Alcántara, Garret Ríos and Fierro Alonso, 2006) and the main economic activity is agriculture, but the arid climate contributes to the extreme poverty, historically associated to the region (SAGARPA, 2003; Instituto Nacional de Lenguas Indígenas (Mexico), 2009; Rossette, 2017).

Wastewater is received from Mexico City through three canals: West Interceptor Tunnel, Central Interceptor Tunnel, and Grand Drainage Canal (Fig. 1b). Subsequently, the water is collected in the Irrigation Districts (003-Tula, 100-Alfajayucan, and 112-Ajacuba), and a dam system that includes Taxhimay, Requena, Endhó, Javier Rojo Gómez, and Vicente Guerrero dams. This dam system is considered the largest in the world (Islas, 2010). Wastewater is partially processed in the wastewater treatment plant (WWTP) in Atotonilco de Tula, Hidalgo (Fig. 1c), with a maximum treatment capacity of 50 m^3^/s. In Hidalgo, five Irrigation Districts distribute 1,513,395,400 m^3^/year of water (CONAGUA, 2019); 97% corresponds to the irrigation districts 003 of Tula, 100 Alfajayucan, and 112 Ajacuba, which provide residual water to the agricultural sector of the Mezquital Valley and belong to the hydrological-administrative region XIII Mexico Valley.

The WWTP normally works at 30% of its capacity because the producers refuse to use the treated water in their irrigation systems. For this reason, this is a health and environmental risk because these wastewaters irrigate more than 80,000 ha of crops (Lesser *et al*., 2018). However, the use of wastewater from Mexico City has had a positive impact on the productivity of the diverse crops grown (Institute for Federalism and Municipal Development (INAFED), 2010). Wastewaters contribute with large amounts of organic matter during decreasing the costs of fertilization. These factors have allowed an increase in land rentability, reaching >1,000 USD per ha per year. In turn, wastewater treatment could directly increase production costs, price of water, and increased needs for fertilizers, which could impact land rentability (Pérez Camarillo, 2002; Pérez, Zacatenco and Martínez, 2006; SIAP, 2018).

### Sample collection

Between September 22^nd^ and October 6^th^ 2020, we collected water samples in eight locations of the Tula River, which represent the main flow of wastewater from Mexico City in the Mezquital Valley, this also included two locations in the Tepeji River and Salado River (Fig. 1c, Table 1). Water samples were also collected from irrigation canals adjacent to agricultural fields as well as soil samples from these agricultural fields (Fig. 1c, Table 1). Produce samples were also collected at three of the agricultural fields: a sample of coriander at site 3, and samples of lettuce at sites 6 and 7. In total, our sampling covered ≈80 km in the Mezquital Valley, and included the municipalities of Tepeji del Río, Atotonilco de Tula, Tula de Allende, Tezontepec de Aldama, Mixquiahuala de Juárez, Tlaxcoapan, Tetepango and Ixmiquilpan (Table 1).

**Table 1.** Water samples collected in Mezquital Valley for quantification of SARS-CoV-2.

| Code | Location | Waterbody | Latitude | Lenght | Description |
| --- | --- | --- | --- | --- | --- |
| RW1 | Tepeji-El Salto Bridge | Tepeji River | 19.91062 | -99.33189 | A mix of water-springs and municipal discharge from localities of Tepeji del Río de Ocampo flows into the Requena Dam, which feeds the Tula River. |
| RW2 | Central Interceptor Tunnel Mouth | Tula River | 19.953149 | -99.297094 | Receives wastewater discharges from the Mexico City Metropolitan Area (MCMA). These are discharged into “El Salto” river, which becomes “Tula” river from this point. |
| RW3 | Cooperativa Cruz Azul City Bridge | Tula River | 19.987822 | -99.323474 | Intermediate location between *Emisor Central* and the city of Tula de Allente. |
| RW4 | Tula de Allende City Bridge | Tula River | 20.05919 | -99.340291 | Intermediate location between *Requena* Dam and *Endhó* Dam |
| RW5 | Tezontepec City Bridge | Tula River | 20.194539 | -99.281055 | Intermediate location between *Endhó* Dam and the convergence of *Salado* river and *Tula* river. |
| RW6 | Mixquiahuala City Bridge | Tula River | 20.240323 | -99.230846 | Location downstream of the convergence of *Salado* river and *Tula* river. |
| RW7 | Mangas Bridge | Salado River | 20.184949 | -99.25571 | *Salado* river; accumulates municipal wastewater and receives wastewater from MCMA *Gran Canal de Desague* |
| RW8 | Ixmiquilpan | Tula River | 20.483365 | -99.22058 | Tula river at Ixmiquilpan; this |
|  | City Bridge |  |  |  | location if 80 km downstream of *Emisor Central* |
| CW1 | Requena Canal | Final Canal | 20.191971 | -99.243117 | Water from the Wastewater Treatment Plant (WWTP). In the literature, the WWTP registers that it treats only 30% of all the flow from the MCMA. However, the CONAGUA assures that it currently treats 40 to 60%. |
| CW2 | Endho Canal | Intermediate Canal | 20.215935 | -99.203465 | Water from the Endhó Dam. Once the Tula River leaves the city of Tula de Allende, it reaches the Endhó Dam. |
| CW3 | Requena Canal | Main Canal | 20.156447 | -99.234083 | Water from the Wastewater Treatment Plant. |
| CW4 | WWTP Canal | Main Canal | 19.963483 | -99.301255 | According to producers, the WWTP only treats less than 50% of the Central Interceptor Tunnel discharge effluent. The treated water is mixed with residual water, and the flow continues through the irrigation canals. |
| CW5 | Fuerza Canal (Tlamaco-Juandho) | Final Canal | 20.087938 | -99.2049 | It is also called Tlamaco-Juandhó. It carries a mixture of water from municipal discharges and wastewater from the WWTP, Tequisquiapan, and Zumpango. |
| CW6 | Alto-Ajacuba Canal | Final Canal | 20.105037 | -99.146827 | The endpoint of irrigation canals, coming from the WWTP and a mixture of municipal discharges and thermal-watering places. This area already belongs to the |
|  |  |  |  |  | Irrigation District 112-Ajacuba. |
| CW7 | Alto Canal | Final Canal | 20.521108 | -99.133735 | Water from the Endhó Dam, passing through the Tula River. |
| CW8 | Xotho Canal | Intermediate Canal | 20.526834 | -99.1095 | Water from the slopes of the Tepatepec, Tezontepec, Mixquiahuala Canals. |

RW; water sample from Tula River. CW; water sample from irrigation canal.

### SARS-CoV-2 quantification in water samples from Tula River

We detected RNA from SARS-CoV-2 in water samples collected in the Tula River, Tepeji River and Salado Rover (Fig. 2). Detection was achieved from the three targets included in the kit (N1-FAM, N2-HEX and N3-TexasRed) in most cases; however, quantification using N2-HEX Ct values and its corresponding standard curve yielded more consistent results, and absolute values were always intermediate to those obtained with N1-FAM and N3-TexasRed (not shown). From these criteria, we use N2-HEX data throughout this study. SARS-CoV-2 RNA was detected in 6 out of 8 water samples from river (Fig. 2). As expected, the highest concentration was found in sample RW2, at the entrance flow of the wastewater treatment plant (WWTP, Fig. 1c and 2), with a concentration of 79 RNA copies per ml; from the sampled locations in the Tula River, this is the closest to the Mexican Valley Metropolitan Area (MVMA, Fig. 1 and 2). This value decreased through the flow of the river (which consists mostly of untreated wastewater) in samples RW3 and RW4, and was undetectable in RW5, downstream of the Endhó Dam. SARS-CoV-2 was also detected at the entrance fluxes of Tepeji River (RW1, 18 copies per ml) and Salado River (RW6, 24 copies per ml), both of which contribute to viral load in the Tula River. Finally, SARS-CoV-2 was not the detected in sample RW8, which was collected at Ixmiquilpan, ≈40 km downstream of sample RW7 (Fig. 2).

**Figure 2.**
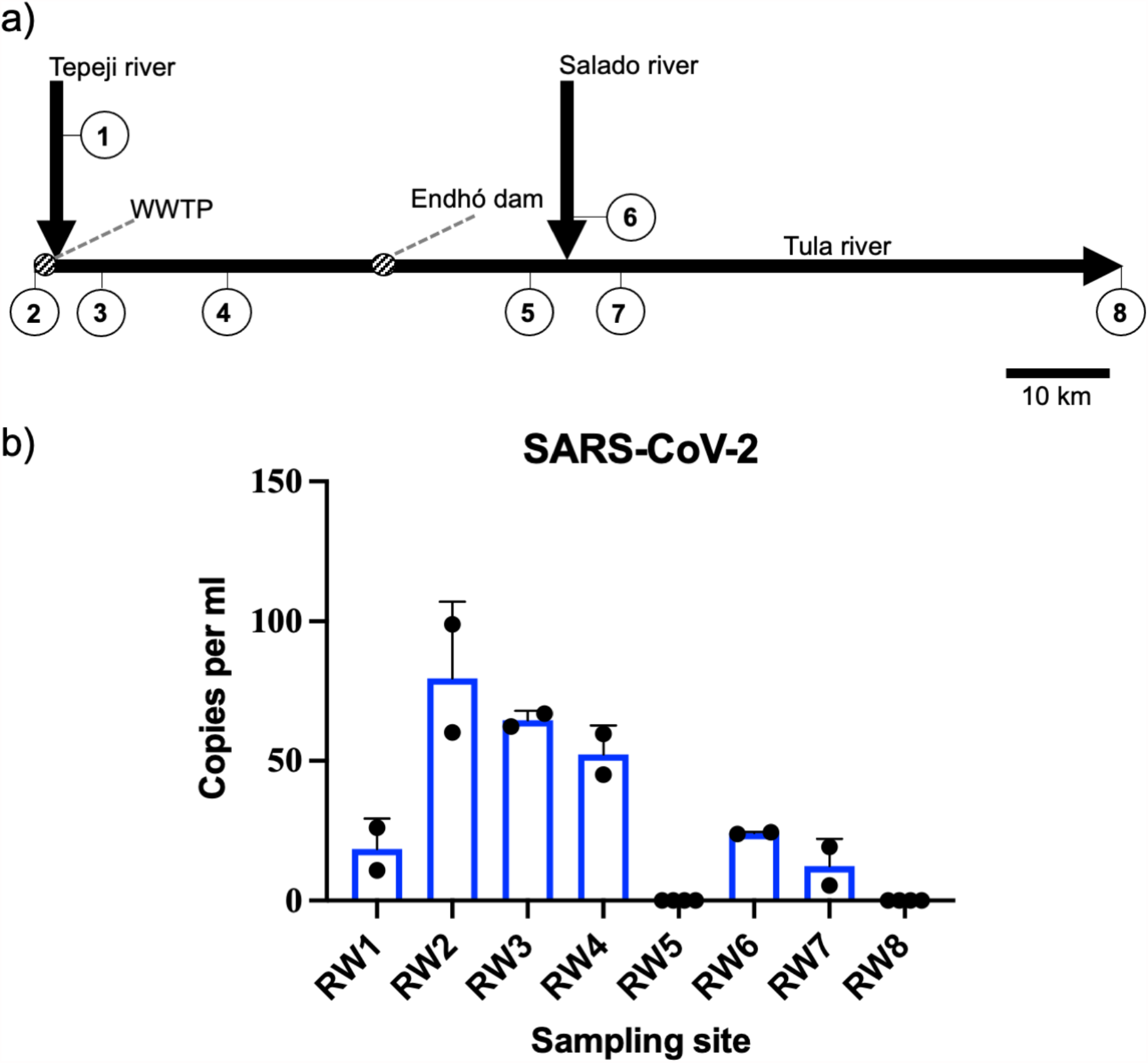
Quantification of SARS-CoV-2 ARN in water samples from the Tula River. a) Schematic representation of distances and fluxes present between the sampling locations at the Tula, Tepeji and Salado River. WWTP, Wastewater treatment plant. b) RT-qPCR results of SARS-CoV-2 quantification in the Tula River.

### SARS CoV-2 RNA in irrigation canals

RNA from SARS-CoV-2 was also detected in water samples collected in irrigation canals throughout the Mezquital Valley. Irrigation canals are mostly fed from treated water, however, untreated wastewaters from municipalities in Hidalgo are also connected to these canals. Since the irrigation canal network in the Mezquital Valley is highly complex and a detailed map is unavailable, we only show the general known source of water in each location (Fig. 3a). RNA From SARS-CoV-2 was detected in 5 out of 8 water samples collected from irrigation canals (Fig. 3b). The highest concentration of 112 copies per ml was found in sample CW1 (Fig. 3b). This location is the closest from MVMA, however, it is reportedly fed solely from wastewater treated in the WWTP (Fig. 3a, table 1). SARS-CoV-2 ARN was also detected in samples CW2, CW4, CW4-II and CW5, all of which are fed from the WWTP, but also from wastewater from municipalities in Hidalgo, and from the Endhó Dam (Fig. 3). SARS-CoV-2 was not detected in samples CW3, CW6 and CW7; notably, irrigation canals at Ixmiquilpan (CW6 and CW7) are fed from freshwater bodies (not wastewater) such as groundwater wells or local spas, while CW3 is reportedly fed from the WWTP and local municipalities (Fig. 3).

**Figure 3.**
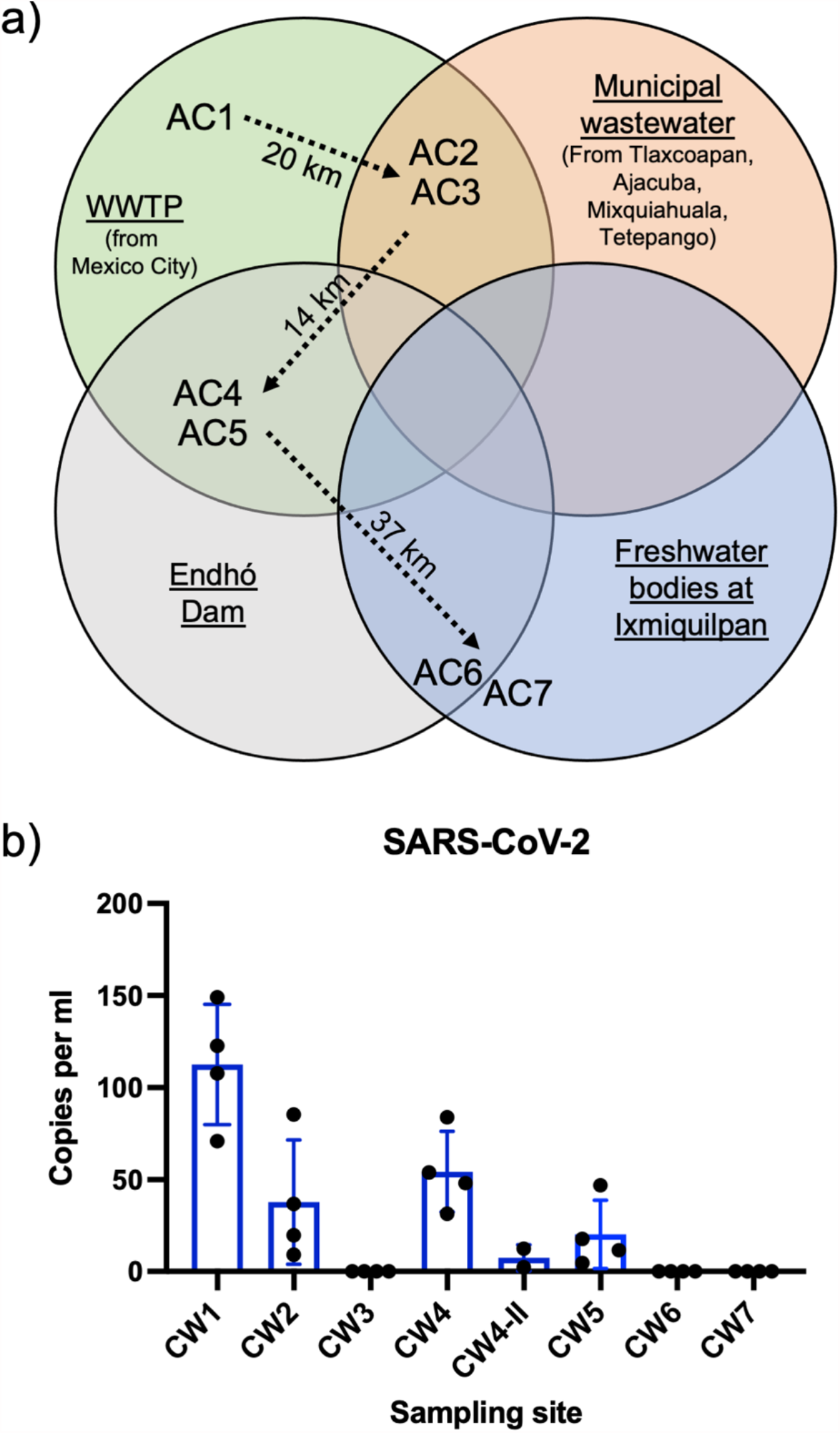
Quantification of SARS-CoV-2 ARN in water from irrigation canals in the Mezquital Valley. a) Schematic representation showing the reported source of water in irrigation canals sampled. b) RT-qPCR results of SARS-CoV-2 quantification in water samples from irrigation canals.

Since SARS-CoV-2 RNA was detected in water samples from irrigation canals, we hypothesized that it may be detectable in agricultural soil from adjacent fields (Fig. 1c, Table 1). However, SARS-CoV-2 was not detected in these samples (Supplementary Table S1 and S2). Similarly, SARS-CoV-2 RNA was not detected in any of the produce samples, which were collected in the field or at the Ixmiquilpan Market (Table 1, Supplementary Table S2). Since the internal control PMMV was detected in all of these samples (Supplementary Table S2), we discard that the presence of PCR inhibitors as the reason for lack of detection, but further studies are needed to confirm the absence of SARS-CoV-2 since viral particles can adsorb to soils and other solids.

### Physicochemical and microbiological analyses of water samples

Results on physicochemical analyses are shown in Supplementary Table S3 and results on microbiological analyses are shown on Supplementary Table S4. Based on standards stablished by Mexican laws, all water samples had values of BOD, TSS and SS below the permitted limits for water used for irrigation (NOM-001-SEMARNAT-1996). Regarding limits for microbiological parameters (NOM-001-ECOL-1996), 5 out of 8 water samples from the Tula River, and 5 out of 8 water samples from irrigation canals, presented total coliform values above the allowed limits for wastewater used for irrigation (Supplementary Table S4).

We assessed if the physicochemical characteristics of wastewater, organic matter and microbial concentration correlated with the concentration of the virus. In samples from the river water, correlation analyses showed that several variables were associated with SARS-CoV-2 RNA concentration. In contrast, analyses using parameters obtained from irrigation canals, showed no significant correlations between SARS-CoV-2 RNA concentration and physicochemical or microbiological variables (Table 2, right). Significant *p* values were obtained for correlation analyses with physicochemical parameters BOD, COD, pH, and SS (Table 2, left), and also for all three microbiological parameters in river samples (Table 2, Supplementary Figure S1). Further inspection of correlation trends revealed correlations with pH and SS could be spurious, since they were highly grouped in discrete values (Supplementary Figure S1), and a higher number of samples should be analyzed to confirm this observations (Table 1, Suplementary Figure S1).

**Table 2.** Correlation of SARS-CoV-2 quantification versus physicochemical and microbiological parameters in water samples.

| Physicochemical parameters |  |  |  |  |  |  |
| --- | --- | --- | --- | --- | --- | --- |
| Parameter | River water samples |  |  | Irrigation canal samples |  |  |
| | $r^2$ | $p$ value | Significance | $r^2$ | $p$ value | Significance |
| BOD | 0.66451416 | 0.00872825 | ** | 0.11417103 | 0.40483251 | - |
| **COD** | 0.94529181 | 7.42E-06 | *** | 0.0181819 | 0.74745041 | - |
| **pH** | 0.68970848 | 0.0064762 | ** | 0.42338299 | 0.06905202 | - |
| **TDS** | 0.24418989 | 0.20131137 | - | 0.3650173 | 0.10036602 | - |
| **SS** | 0.86636172 | 0.00024926 | *** | 0.00302837 | 0.89594555 | - |
| **TSS** | 0.26441269 | 0.18013422 | - | 0.01181227 | 0.79562296 | - |
| **TS** | 0.17469837 | 0.29241941 | - | 0.35416452 | 0.10724613 | - |

Microbiological parameters
| Parameter | River water samples |  |  | Irrigation canal samples |  |  |
| --- | --- | --- | --- | --- | --- | --- |
|  | r<sup>2</sup> | p value | Significance | r<sup>2</sup> | p value | Significance |
| **TC** | 0.82551851 | 0.00070422 | *** | 0.21502632 | 0.23573855 | - |
| **FC** | 0.80436747 | 0.00109741 | *** | 0.35713184 | 0.10532899 | - |
| ***E. coli*** | 0.80436747 | 0.00109741 | *** | 0.40315677 | 0.07888845 | - |

BOD, Biochemical oxygen demand; COD, chemical oxygen demand; pH, Hydrogen potential, TDS, total dissolved solids; SS, suspended solids, total suspended solids; TSS, total suspended solids; TS, total solids; TC, total coliforms; FC, fecal coliforms; \*\*, $p < 0.01$ ; \*\*\*, $p < 0.005$ .

COD and BOD are a measurement of oxygen required for chemical oxidation of organic material in water, and the Biodegradability index (COD/BOD ratio) is an indicator of the non-biodegradable organic-chemicals. We assesed if the Biodegradability index correlates with the virus concentration of the water system (Abdalla and Hammam, 2014). Interestingly, the trend is the same than the virus concentration (Supplementary Figure S2).

### Modelling the geographic dispersion of SARS-CoV-2 in the Mezquital Valley

Using data for SARS-CoV-2 concentration in the Tula River, we sought to adapt a model that describes viral dispersion in the wastewater bulk. Our model only considers the dispersion of the virus in water and the modification of the flux in the mainstream. The dispersion coefficient was calculated through least square optimization to the experimental data (Suplementary File 1) (Molugaram et al., 2017). The concentration in the initial sampling site RW2 (Figure 1c.) was normalized to 1 (Ratio of Concentration C/initial Concentration Ci); since it was the maximun value, the experimental data were normalized to this value. Additionally, we modeled two entrances at the points RW2 and RW6, as a result the dispersion coefficient increases at these points. The model fitted to the experimental data along the Tula River (Figure 4a). Therefore, with only three variables, dispersion coefficient (*D*), amount of water (Θ) and a distance function of *D* [*D*(x)], we could define the dispersion of the virus in the Tula River. Finally, we used QGIS3 to obtain a geographic representation of our model and the experimental data (Figure 4b), which show the general trends of SARS-CoV-2 dispersion in the Tula River.

**Figure 4.**
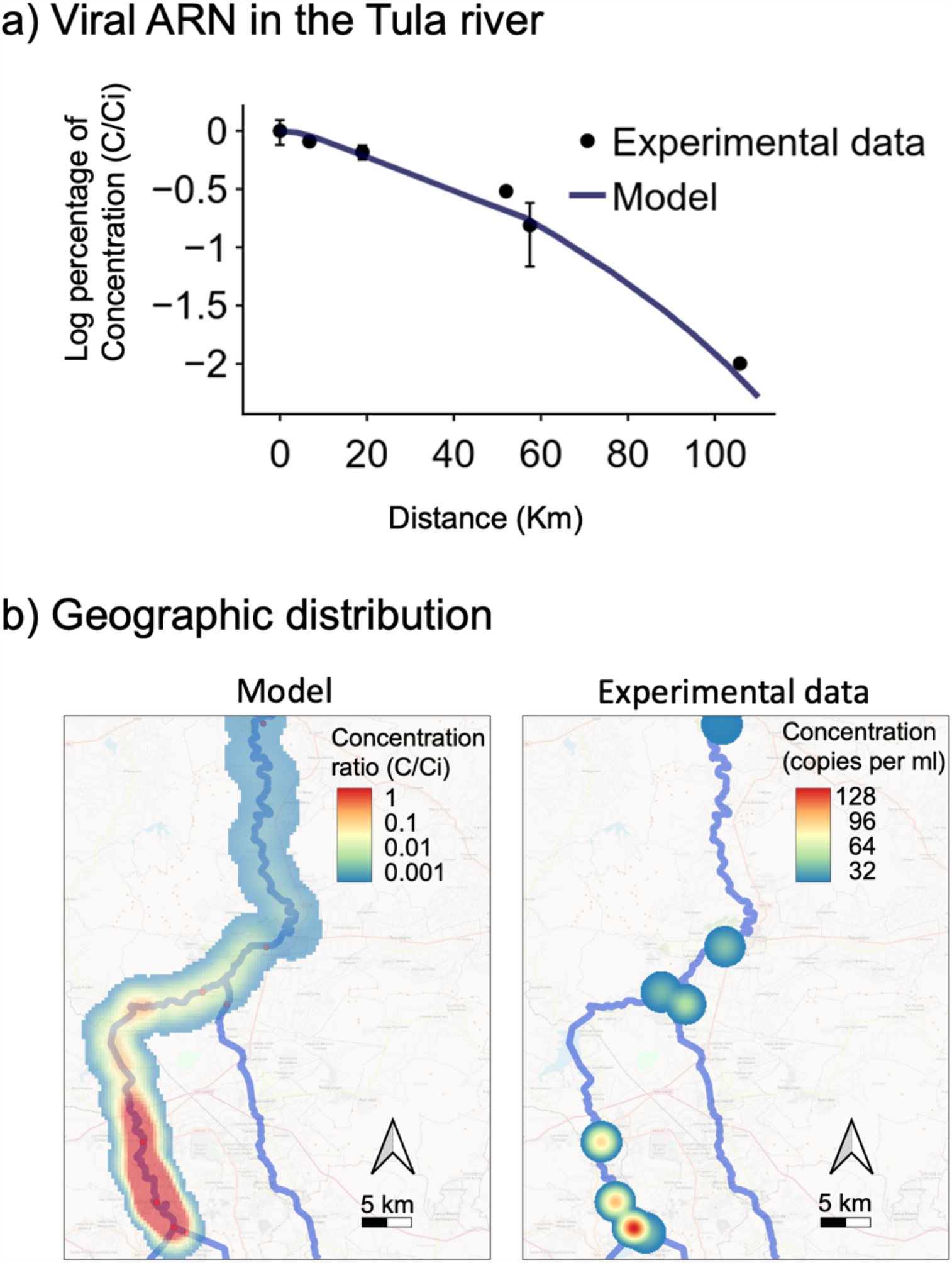
The transport model generated for SARS-CoV-2 dispersion in the Tula River, fits the data obtained experimentally. a) Graphic representation of viral concentration in the Tula River, comparing the experimental data and the transport model obtained in this study. b) Map representation of the geographic dispersion of SARS-COV-2 presence in the Tula River, comparing the transport model (left) and the experimental data (right).

## Discussion

In this work, we quantified SARS-CoV-2 ARN in wastewaters from Mexico City, which are distributed throughout the Tula River, Salado River and irrigation canals and are used for irrigation of crops in the Mezquital Valley. Notably, we detected the virus in geographically representative water samples from both rivers and the irrigation canals. With our data, we generated a viral dispersion model which can be used to assess concentration of SARS-CoV-2 to other river systems if the dispersion coefficient (*D*), amount of water (Θ) and a distance function of *D* [*D*(x)] of the river are fitted.

Samples from both the Tula River and irrigation canals followed the same general trend of presenting the highest concentration in the sites located near the WWTP. Viral concentration decreased in the remaining sites and was undetectable in the northernmost sites, located near the town of Ixmiquilpan. This result indicates that the most important source of SARS-CoV-2 ARN in these water systems is the wastewater from Mexican City Valley, and that the viral particles tend to degrade or dilute along the flow of water through the Mezquital Valley. However, we do not discard that municipalities in the Mezquital Valley contribute to viral load and fecal contamination of Tula River, Salado River and the irrigation canals.

Our results showed that the SARS-CoV-2 virus was not detected on soil samples. While this could be related to PCR inhibitors in the samples, we found that the PMMV virus was detected and hence discarded this hypothesis. Virus associate with soil and other particulate matter and both adsorption and detachment of virus depends on temperature, moisture, pH, and the physicochemical characteristics of the virus capsid surface and the particles. Similarly, the microbes and organic matter may have a protective effect on virus (AzadpourKeeley, Faulkner and Chen, 2003). Further studies are needed to asses if lack of detection of SARS-CoV-2 is related to degradation or dispersion of the virus, or alternatively, if viral particles adsorb in the soil matrix, which would make our ARN extraction method not appropriate.

We found that physicochemical variables COD and BOD and microbiological variables showed significant correlations with SARS-CoV-2 quantification, but only in samples from the Tula River. However, these observations should be confirmed with measurements in a higher number of samples. This result suggests that in the river, anthropological sources from a single source of contamination predominate, i.e. the MVMA. However, in irrigation canals – where no correlations were found – may receive other sources of contamination (from agriculture, livestock, or industry) (Guédron *et al*., 2014; Lesser *et al*., 2018). Furthermore, our correlation analyses suggest that BOD, COD, and the Biodegradability Indec (BOD/COD), as well as fecal coliforms may be good indicators of SARS-CoV-2 ARN concentration in municipal wastewaters.

The model proposed to explain the dispersion of SARS-CoV-2 virus along the Tula River, as well as the experimental data, indicate that the viral concentration decreases from south to north, with an initial dispersion point at the entrance of the Tula River at the WWTP and a minimal contribution from the communities along the Mezquital Valley. Our model predicts viral dispersion in the Mezquital Valley, and may be useful for selection sampling locations prior to temporal monitoring to quantify the evolution of SARS-CoV-2 (or other virus) epidemics in populations. Previously available models for viral dispersion in environmental matrices (Bivins *et al*., 2020; Farkas *et al*., 2020; Kitajima *et al*., 2020) are useful for detailed dissection of variables governing dispersion patterns of viral particles in at a smaller scale in controlled environments. Our model is, to our knowledge, the first to allow prediction of viral dispersion in linear water bodies at a geographic scale. This model could be extrapolated to other rivers, streams, and canals.

One of the main purposes of this work was to assess the possible risk of environmental transmission of SARS-CoV-2 through environmental matrices. Indeed, some recent reports suggest/show that the virus may be infective (Kitajima e*t al*., 2020), and hence, direct contact with water bodies in the Mezquital Valley may represent a risk of transmission, especially for farmers during flooding irrigation (Lüneberg *et al*., 2018). However, this work does not seek to eliminate wastewater usage in the Mezquital Valley, as this practice is one of the key factors that allowed the economic development of the region. In fact, the Mezquital Valley is considered a study model for the agricultural development of arid and semi-arid rural regions (Anderson, 2020; Bonvehi Rosich and Seth Denizen, 2021; Seth Denizen, 2021). We suggest that our work should be considered for the creation and modification of practices and policies related to water treatment (Belhadi *et al*., 2020), pathogen monitoring, agricultural practices, and safety measures for farmers in the Mezquital Valley.

## Data Availability

Additional data on mathematical model is available in the link provided.

https://nbviewer.jupyter.org/github/yaxastro3000/COVID_CIAD_URH/blob/c65ac45af14736023e94eb087aea8d541a7dac68/MODEL_COVID_CIAD_URH.ipynb

## 5. Acknowledgements

This work was supported by CONACyT, Mexico, grant 312014 for JR. This work was possible thanks to the collaboration of local farmers in the state of Hidalgo, who allowed us to collect samples in their fields. The authors thank Gabriela Rivas (UPFIM) and Ana Alcalá (IBT UNAM), Laura Palomares (IBT), Oscar Monroy (UAMI) and Jesús Hernández (CIAD Hermosillo) for advice and technical assistance. We thank personnel from governmental institutions in the state of Hidalgo: Jaime Ortega (SEDAGROH), Sergio Guzman (CONAGUA) and José Alonso Huerta (CITNOVA), for their valuable contribution in the stages of planning, sampling, and discussion of results.

## Supplementary material captions

A separate supplementary material PDF file is available.

**Supplementary file 1**. Code details, boundary conditions and description of the process for adjusting the transport model of virus in interfaces to an ordinary differential equation that describes the dispersion of SARS-CoV-2 in the Tula River. https://nbviewer.jupyter.org/github/yaxastro3000/COVID_CIAD_URH/blob/c65ac45af14736023e94eb087aea8d541a7dac68/MODEL_COVID_CIAD_URH.ipynb

## Supplementary tables

**Supplementary Table S1**. Soil samples collected for this study.

**Supplementary Table S2**. Ct values obtained from RT-qPCR detection of SARS-CoV-2 and PMMV in viral nucleic acids from soil and produce samples

**Supplementary Table S3**. Physicochemical parameters of water samples from Tula River (RW) and irrigation canals (CW)

**Supplementary Table S4**. Microbiological analyses of water and soil sample

## Supplementary figures

**Supplementary Figure S1**. Correlations between SARS-CoV-2 and physicochemical variables (top) and between SARS-CoV-2 and microbiological variables (bottom) in water samples.

**Supplementary Figure S2**. Biodegradability index (COD/BOD) measured in Tula River, Tepeji River and Salado River, shows trends similar to those od SARS-CoV-2 concentration. Each label represents the sample location indicated in figure 1c.

