## Supplemental Material for "SARS-CoV-2 in wastewater from Mexico City used for irrigation in the Mezquital Valley: quantification and modelling of geographic dispersion"

[1]Conacyt-Unidad Regional Hidalgo, Centro de Investigación en Alimentación y Desarrollo. Ciudad del Conocimiento y la Cultura de Hidalgo, Blvd. Santa Catarina S/N, San Agustín Tlaxiaca, Hidalgo, México, 42163

[2]Unidad Regional Hidalgo. Centro de Investigación en Alimentación y Desarrollo. Ciudad del Conocimiento y la Cultura de Hidalgo, Blvd. Santa Catarina S/N, San Agustín Tlaxiaca, Hidalgo, México, 42163.

[3]Instituto Tecnológico de Celaya. Antonio García Cubas 600, Fovissste, Celaya, Gto., 38010.

[4]Universidad Tecnológica de Querétaro. Av. Pie de la Cuesta 2501, Nacional, Santiago de Querétaro, Qro., 76148.

**Supplementary file 1**. Code details, boundary conditions and description of the process for adjusting the transport model of virus in interfaces to an ordinary differential equation that describes the dispersion of SARS-CoV-2 in the Tula River.
https://nbviewer.jupyter.org/github/yaxastro3000/COVID_CIAD_URH/blob/c65ac45af1473602
3e94eb087aea8d541a7dac68/MODEL_COVID_CIAD_URH.ipynb

**Supplementary tables**

**Supplementary Table S1.** Soil samples collected for this study.

| Sample | Date | Municipality | Coordinates | Days since irrigation |
|---|---|---|---|---|
| S1 | 29/09/2020 | El Salto | 19.963483, -99.301255 | 14 |
| S2 | 29/09/2020 | Tlaxcoapan | 20.090066, -99.215945 | 10 |
| S3 | 29/09/2020 | Tetepango | 20.102991, -99.149837 | 14 |
| S4 | 23/09/2020 | Mangas | 20.1909989, -99.2430968 | 4 |
| S5 | 23/09/2020 | Palmillas | 20.215935, -99.203465 | 7 |
| S6 | 06/10/2020 | Debodhé | 20.521108, -99.133735 | 4 |
| S7 | 06/10/2020 | Debodhé - Cerro | 20.526567, -99.110138 | 4 |

**Supplementary Table S2.** Ct values obtained from RT-qPCR detection of SARS-CoV-2 and PMMV in viral nucleic acids from soil and produce samples

| Sample | SARS-CoV-2 (Ct) | | | PMMV (Ct) |
|---|---|---|---|---|
| | N1-FAM | N2-HEX | N3-TexasRed | |
| S1 | ND | ND | ND | 30.264921 |
| S2 | ND | ND | ND | 31.091442 |
| S3 | ND | ND | ND | 26.787857 |
| S4 | ND | ND | ND | 29.487373 |
| S5 | ND | ND | ND | 28.38734 |
| S6 | ND | ND | ND | 31.776516 |
| S7 | ND | ND | ND | 31.768978 |
| Milpa | ND | ND | ND | 37.909 |
| Natural landscape | ND | ND | ND | ND |
| Coriander 3 | ND | ND | ND | 38.9628 |
| Lettuce 6 | ND | ND | ND | 36.6487 |
| Lettuce 7 | ND | ND | ND | 36.3314 |
| Broccoli M | ND | ND | ND | 32.7974 |
| Cauliflower M | ND | ND | ND | 26.0486 |
| Squash M | ND | ND | ND | 30.8299 |
| Positive control | 23.8252 | 21.9638 | 27.3536 | NA |
| Negative control | ND | ND | ND | ND |

ND, not detected; NA, not determined. Numbers in produce samples correspond to locations of fields from Figure 1. M, samples collected at the Ixmiquilpan market.

**Supplementary Table S3. Physicochemical parameters of water samples from Tula river (RW) and irrigation canals (CW)**

| Sample | BOD (mg/L) | COD (mg/L) | pH | TDS (mg/L) | SS (ml/L) | TSS (mg/L) | TS (mg/mL) |
|---|---|---|---|---|---|---|---|
| **RW1** | 5.5 | 122 | 8.1 | 345.3 | 0 | 1.8 | 347.1 |
| **RW 2** | 19.9 | 244 | 7.9 | 693 | 1 | 41 | 734 |
| **RW 3** | 21.8 | 219.6 | 8.1 | 580 | 1 | 41 | 621 |
| **RW 4** | 21.5 | 146.4 | 8.1 | 524 | 1 | 126 | 650 |
| **RW 5** | 8.4 | 30 | 8.6 | 858.6 | 0 | 15 | 873.2 |
| **RW 6** | 12 | 105 | 8.1 | 823.7 | 0.1 | 44.25 | 863 |
| **RW 7** | 13 | 60 | 8.6 | 967.8 | 0 | 8,4 | 976.2 |
| **RW8** | 12.9 | 27.1 | 8.6 | 1,016.00 | 0.1 | 10.6 | 1,027.00 |
| **CW1** | 35.9 | 120 | 7.7 | 735 | 0 | 49.6 | 784.6 |
| **CW2** | 34 | 210 | 7.7 | 711.4 | 0.8 | 91.5 | 802.9 |
| **CW3** | 19 | 255 | 8.1 | 1039.3 | 0.3 | 145.5 | 1184.8 |
| **CW4** | 20.6 | 195.2 | 8.3 | 765.3 | 0.6 | 67 | 832.3 |
| **CW4-II** | 22.7 | 219.6 | 8.3 | 770 | 0.4 | 25 | 795 |
| **CW5** | 21.5 | 60 | 8.2 | 734.6 | 0 | 16.53 | 751.1 |
| **CW6** | 42.6 | 10.2 | 9 | 1189 | 0 | 4 | 1193 |
| **CW7** | 15.5 | 4.5 | 9 | 1252.4 | 0 | 4.7 | 1257.2 |

BOD, Biochemical oxigen demand; COD, chemical oxigen demand; pH, Hydrogen potential, TDS, total dissolved solids; SS, suspended solids, total suspended solids; TSS, total suspended solids; TS, total solids.

**Supplementary Table S4.** Microbiological analyses of water and soil samples

| **Sample** | ***Salmonella spp.**** | **TC (NMP/100mL)** | **FC (NMP/100mL)** | ***E. coli* (NMP/100mL)** |
|---|---|---|---|---|
| RW1* | Detected | >110 000 | 110 000** | 110 000 |
| RW2* | Detected | 408 000 000 | 99 500 000** | 99 500 000 |
| RW3* | Detected | >11 500 000 | 11 500 000** | 11 500 000 |
| RW4* | Detected | >11 500 000 | 171 000** | 171 000 |
| RW5 | Detected | >11 000 | 360 | 360 |
| RW6 | Detected | >11 000 | 300 | 300 |
| RW7* | Detected | >11 000 | >11 000** | >11 000 |
| RW8 | Detected | 46 000 | 1 500 | 1 500 |
| CW1* | Detected | 9 200 000 | 9 200 000** | 9 200 000 |
| CW2* | Detected | 23 000 000 | 3 600 000** | 3 600 000 |
| CW3 | Detected | 9 200 000 | <300 000** | <300 000 |
| CW4* | Detected | >11 500 000 | 11 500 000** | 11 500 000 |
| CW4-II* | Detected | 11 500 000 | 11 500 000** | 4 080 000 |
| CW5* | Detected | >11 000 | >11 000** | >11 000 |
| CW6 | Detected | 2 700 | 740 | 740 |
| CW7 | Detected | 15 000 | 740 | 740 |

TC, total coliforms; FC, fecal coliforms; MPN, most probable number.

*PCR detection from 25 g or 25 ml of sample

**Above the limits of (fecal coliforms) stablished in NOM-001-ECOL-1996.

### Supplementary figures

**Supplementary Figure S1.** Statistically significant correlations found between SARS-CoV-2 and physicochemical variables (top) and between SARS-CoV-2 and microbiological variables (bottom) in water samples from the Tula River.

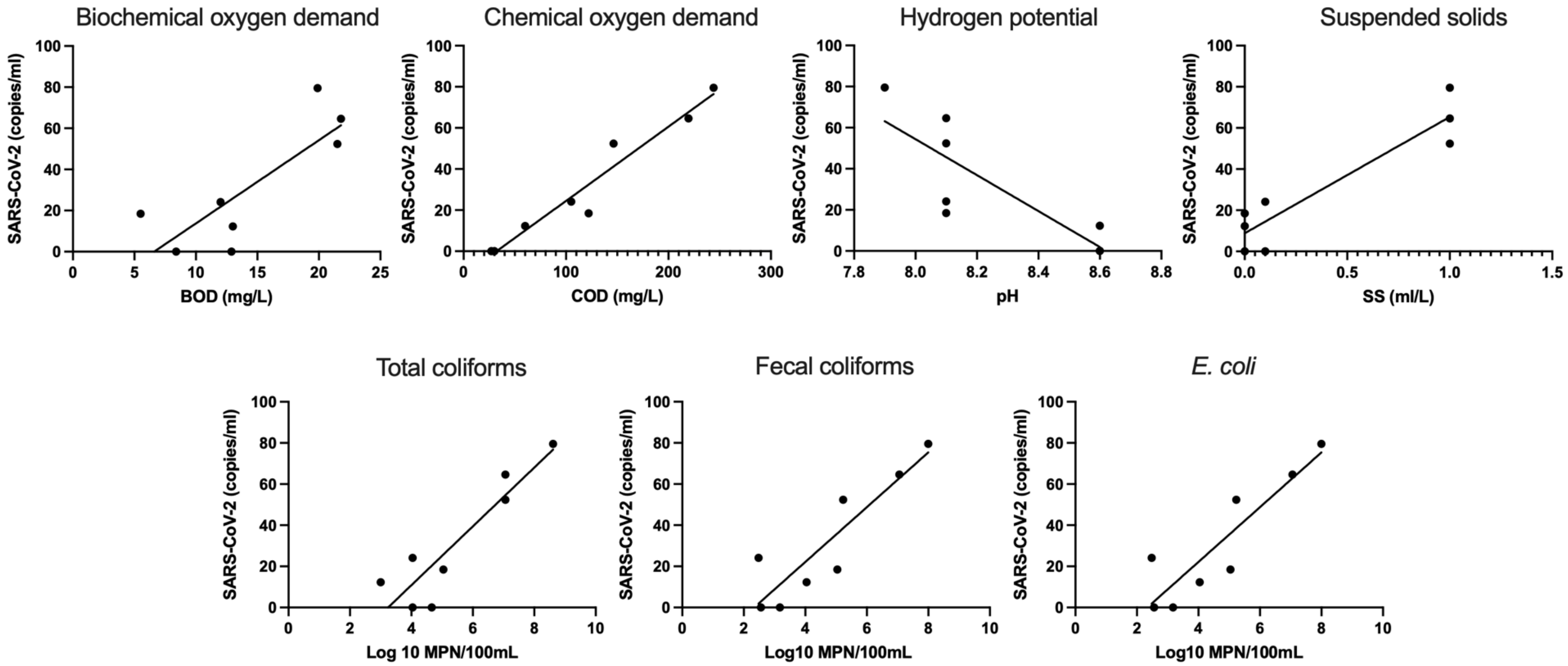

**Supplementary Figure S2.** Biodegradability index (COD/BOD) measured in Tula River, Tepeji River and Salado River, shows trends similar to those od SARS-CoV-2 concentration. Each label represents the sample location indicated in figure 1c.

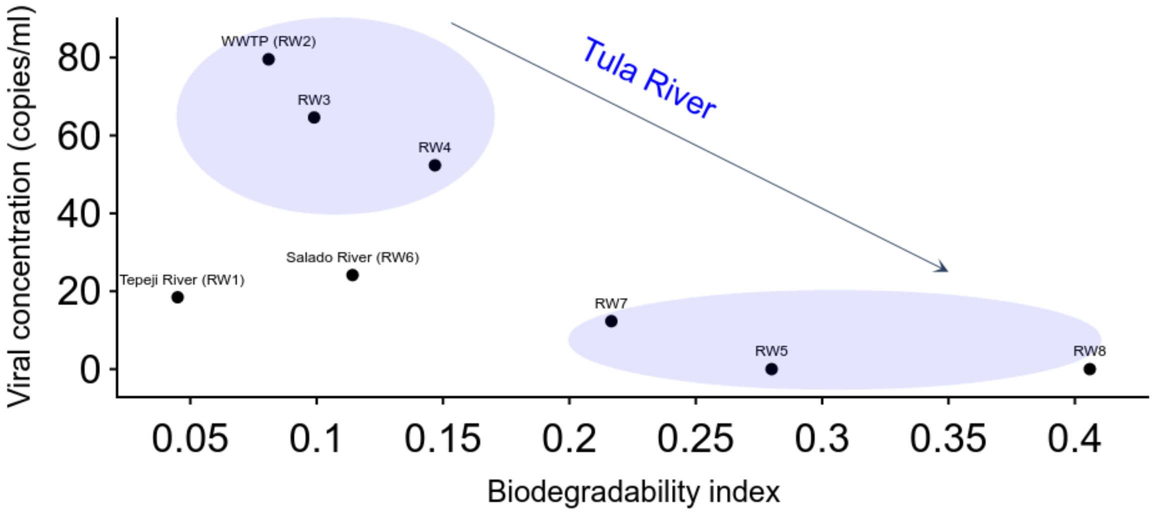
